# A brief report: *de novo* copy number variants in children with attention deficit hyperactivity disorder

**DOI:** 10.1101/2019.12.12.19014555

**Authors:** Joanna Martin, Grace Hosking, Megan Wadon, Sharifah Shameem Agha, Kate Langley, Elliott Rees, Michael J Owen, Michael O’Donovan, George Kirov, Anita Thapar

## Abstract

**Background:** Recent case-control genetic studies of attention deficit hyperactivity disorder (ADHD) have implicated common and rare genetic risk alleles, highlighting the polygenic and complex aetiology of this neurodevelopmental disorder. Studies of other neurodevelopmental disorders, such as autism spectrum disorder (ASD), Tourette disorder, developmental delay/intellectual disability, and schizophrenia indicate that identification of specific risk alleles and additional insights into disorder biology can be gained by studying non-inherited *de novo* variation. In this study, we aimed to identify large *de novo* copy number variants (CNVs) in children with ADHD.

**Methods:** Children with a confirmed diagnosis of ADHD and their parents were genotyped and included in this sample. We used PennCNV to call large (>200kb) CNVs and identified those calls that were present in the proband and absent in both biological parents.

**Results:** In 305 parent-offspring trios, we detected 14 *de novo* CNVs in 13 probands, giving a mutation rate of 4.6% and a per individual rate of 4.3%. This rate is higher than published reports in controls and similar to those observed for ASD, schizophrenia and Tourette disorder. We also identified *de novo* mutations at 4 genomic loci (15q13.1-13.2 duplication, 16p13.11 duplication, 16p12.2 deletion and 22q11.21 duplication) that have previously been implicated in other neurodevelopmental disorders, two of which (16p13.11 and 22q11.21) have also been implicated in case-control ADHD studies.

**Conclusions:** Our study complements ADHD case-control genomic analyses and demonstrates the need for larger parent-offspring trio genetic studies to gain further insights into the complex aetiology of ADHD.

## Introduction

Attention deficit hyperactivity disorder (ADHD) is a highly heritable (∼70%), common and impairing neurodevelopmental disorder with a complex genetic architecture^1^. Recent case-control genome-wide studies point to the involvement of thousands of relatively common single nucleotide polymorphisms (SNPs)^2^, very rare protein-truncating sequence variants and likely pathogenic missense mutations^3^, as well as large, rare copy number variants (CNVs)^4–6^. Although all of these classes of variant are associated with ADHD risk, identifying specific risk alleles is an important next step. Studies of *de novo* mutations using parent-offspring trios provide an especially powerful approach to gene discovery because the background rate in unaffected individuals is low, and therefore if an elevated rate in cases is demonstrable, such mutations are likely pathogenic and can provide important insights into disease biology^7^.

*De novo* likely-damaging mutations, that include CNVs which intersect genes, have been identified for other neurodevelopmental disorders, such as autism spectrum disorder (ASD), Tourette disorder, developmental delay/intellectual disability (DD/ID), and schizophrenia^7–12^. Individuals with ASD have a rate of *de novo* CNVs that is between 3- and 5-fold higher than in unaffected siblings or control individuals^13^. Individuals with simplex Tourette disorder have a similar rate of *de novo* CNVs to probands with ASD^14^ as do individuals with schizophrenia^7^. Of these neurodevelopmental disorders, the rate of *de novo* CNVs is highest in individuals with DD/ID, at approximately 10%^15^.

Trio-based genetic studies of ADHD are lacking. There has been only one small study and it reported a rate of *de novo* CNVs (1.7% or 3 probands out of N=173) that was only slightly elevated above previous reports of *de novo* rates in controls^16^. In this study, we set out to characterise large *de novo* CNVs in parent-offspring trios of probands diagnosed clinically with ADHD.

## Method

### Sample

Children with ADHD, aged 5 to 17 years old, were recruited from UK child and adolescent mental health and paediatric clinics. ADHD diagnoses based on DSM-IV or DSM-III-R were confirmed via a semi-structured research diagnostic interview (the Child and Adolescent Psychiatric Assessment^17^) by trained, supervised psychologists. Full-scale IQ was assessed using the WISC-IV^18^. A proportion of the probands in the sample (N=136 or 44.6%) have been analysed in previous case-control CNV studies^4,5^. Approval for the study was obtained from the North West England and Wales Multicentre Research Ethics Committees. Written informed consent to participate was obtained from parents and children aged 16 years and older and assent was gained from children under 16 years of age.

### Genotyping, CNV calling and quality control (QC)

DNA was obtained from blood and Oragene (saliva) samples for ADHD probands and both biological parents. N=424 ADHD parent-offspring trios were genotyped using a custom version of the Illumina PsychChip. Only samples with >95% call rate and those that passed a sex check were retained. Only complete trios that passed QC and an additional Mendel check in PLINK (to confirm biological relatedness) were used for CNV calling.

CNVs were called using PennCNV^19^ and nearby calls were merged if the CNVs were separated by less than 50% of their combined length. CNVs were removed if they were called using <10 probes, had size <50kb, probe density <20kb/probe or a PennCNV confidence score<10. Samples were removed if they had LRR-SD>0.3, BAF-drift>0.01 or WF>0.05. Additionally, we applied further, more stringent criteria in ADHD probands and excluded individuals with extreme scores on QC metrics (>4SD than the sample mean) as follows: if they had an apparently excessive CNV load (NSEG>50.8 or KB>16593) or poor probe variance (BAF-SD>0.063). These additional steps were performed to ensure that potential *de novo* CNVs would be of high quality, while being conservative and not removing a whole trio based on reduced data quality in just one parent. N=305 complete trios passed all of the above QC and were kept for analyses. After these steps, only CNVs of size >200kb were retained for analysis, to ensure accuracy of calls.

CNVs were then annotated as ‘transmitted’, ‘non-transmitted’, or ‘likely *de novo*’ based on presence or absence of a call made in either parent and/or proband. All CNVs that were flagged as likely *de novo* were visually examined in the offspring and both confirmed biological parents and only clear-cut, un-ambiguous *de novo* calls were retained for analyses, minimising the false positive rate at the expense of potentially missing real calls. We did not perform validation, as the traces were definitive (see the Supplementary Figures).

We examined whether the loci disrupted by the *de novo* CNVs had previously been implicated in case-control common variant^2^ and rare CNV studies^4–6^ of ADHD. Next we compared the list of *de novo* loci with rare CNV loci implicated in other neurodevelopmental disorders, using previously published studies of ASD^8^, DD/ID^20^, schizophrenia^21,22^, and Tourette disorder^14^. We also annotated each *de novo* CNV locus with genes and examined if any of these genes have been implicated in ASD, ADHD or DD/ID, based on studies of loss-of-function exonic mutations and deleterious missense variation^3,9,23^. A number of regions and genes overlapped across disorders. The merged lists of neurodevelopmental genetic risks consisted of 96 unique CNV loci and 197 unique genes. All loci were converted to genome build hg19. Overlap was defined as a CNV overlapping at least 50% of a previously implicated region or a CNV affecting the coding sequence of a gene in the list of genes of interest.

## Results

In the N=305 trios (of which, 36 or 11.8% were female probands) that passed QC, we detected a total of 14 *de novo* CNVs (N=9 that were >500kb and N=5 that were between 200-500kb) in 13 ADHD probands, with one individual having two CNVs that were >500kb. Two of the *de novo* CNV carriers were female and the rest were male. The overall mutation rate was 4.6% (14/305) and the per individual rate was 4.3% (13/305). Although we did not have population controls available in this study, a recent study of control trios using similar methodology reported a *de novo* mutation rate of 1.0% for CNVs>200kb^24^; thus the rate in ADHD was over four times higher than this comparable control rate. The mean IQ of *de novo* CNV carriers was 83.6 (SD=13.0) and the mean IQ of the rest of the sample was 85.2 (SD=13.5), with 3 (25.0%) of the *de novo* CNV carriers (with non-missing IQ) having comorbid intellectual disability (ID; IQ<70), and 26 (9.6%) of the rest of the sample having comorbid ID. The group of *de novo* CNV carriers was too small to perform a statistical comparison.

Table 1 summarises the identified CNVs, annotated with affected genes. None of these genes overlapped with genes implicated by a recent ADHD genome-wide association study (GWAS)^2^ or the list of genes implicated by recent sequencing studies of ASD, ADHD, and DD/ID^3,9,23^. However, four of the *de novo* CNVs have previously been robustly implicated in other neurodevelopmental disorders^8,14,20–22^, two of which have also been implicated previously in case-control CNV studies of ADHD^4–6^, as summarised in Table 2. In addition, duplications at 16p13.11^25^ and a *de novo* duplication at 15q13.1^26^ have also previously been observed in people with schizophrenia. Given that a proportion of the current sample have previously been included in published case-control CNV studies^4,5^, we determined that eight of the *de novo* loci in our study overlap at least partially (≥50% overlap) with a previously reported CNV in a person with ADHD, with six of these eight *de novo* CNV carriers having taken part in the previous case-control studies. On the other hand, six of the loci in our study (4p16.3, 10q11.22-q11.23, 10q21.3, 10q22.2, 15q26.3, and 16p13.3) are in novel regions and have not been previously reported in individuals diagnosed with ADHD in these large case-control studies.

**Table 1:**
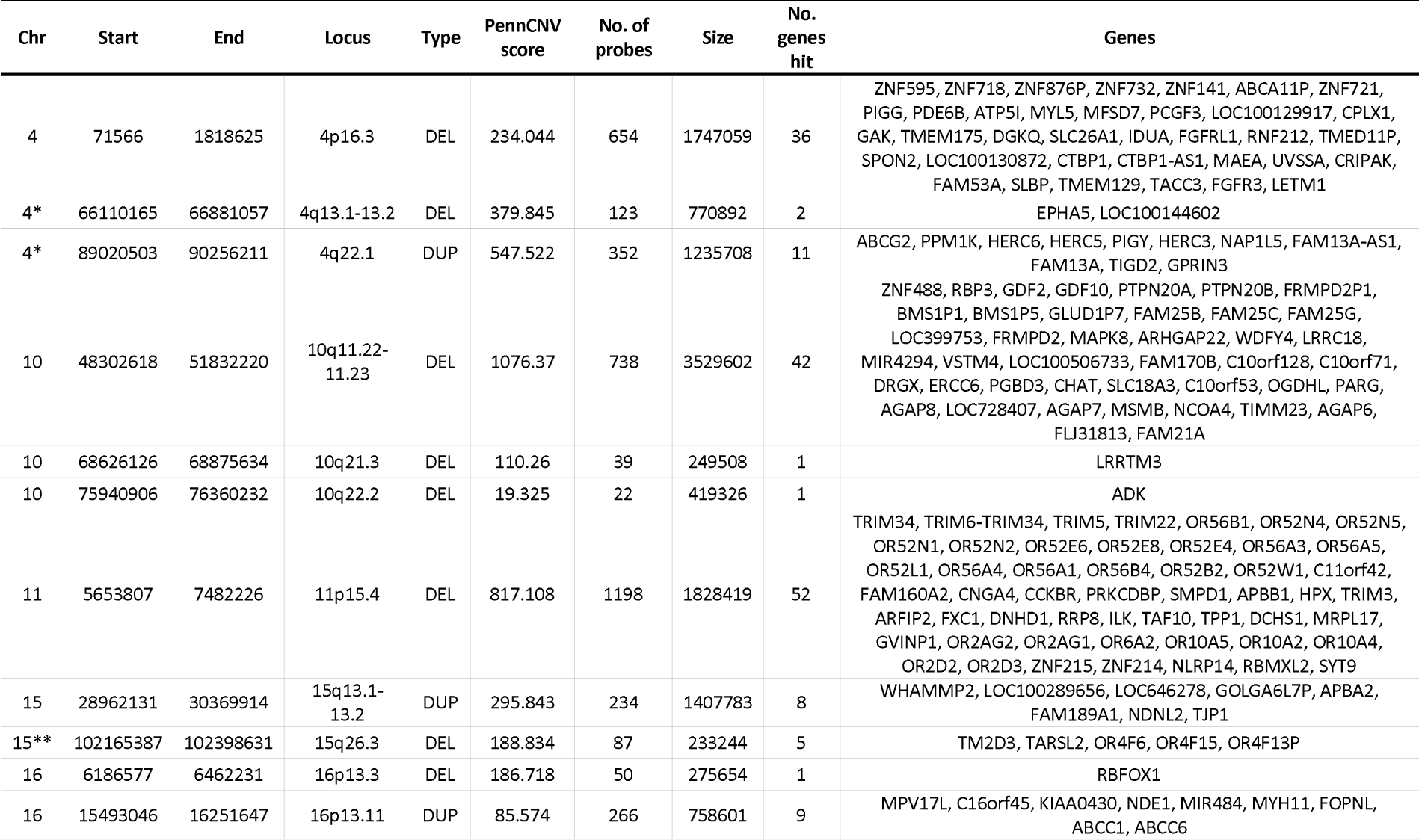
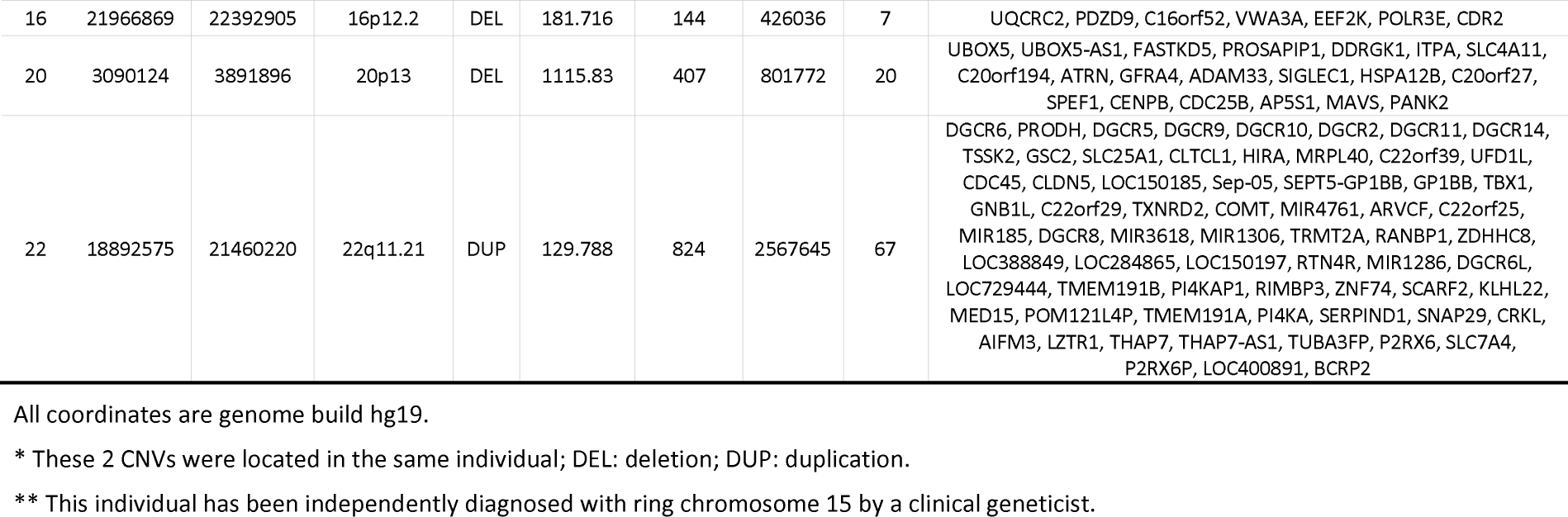
A summary of the identified *de novo* CNVs

**Table 2:** Identified *de novo* CNV regions that have previously been robustly implicated in ADHD and other neurodevelopmental disorders

| CNV | CNV locus previously implicated in ADHD or another disorder |  |  |  |  |
| --- | --- | --- | --- | --- | --- |
|  | ADHD | ASD | DD/ID | SCZ | TD |
| 15:28962131-30369914 (DUP) |  |  |  |  | DUP |
| 16:15493046-16251647 (DUP) | DUP |  | DEL/DUP | DUP |  |
| 16:21966869-22392905 (DEL) |  |  | DEL | DEL |  |
| 22:18892575-21460220 (DUP) | DEL/DUP | DEL/DUP | DEL/DUP | DEL | DUP |

ASD: autism spectrum disorder; DD/ID: developmental delay/intellectual disability; SCZ: schizophrenia; TD: Tourette disorder

## Discussion

In this study, we present observations on *de novo* CNVs from the largest published trio-based study of ADHD to date. The overall mutation rate for *de novo* CNV carriers was 4.6%, which is a similar rate to that observed for two other DSM-5 childhood neurodevelopmental disorders: ASD and Tourette disorder^13,14^, and somewhat higher than observed for schizophrenia^7^. Notably, the rate we observed is substantially higher than that which has been reported for controls^24^ and the previous and only trio-based study of ADHD that observed an overall rate of 1.7%^16^. Aside from the lower power of this previous, smaller study, one potential explanation for the difference is that, unlike Lionel and colleagues^16^, we did not exclude those with an IQ<70. However, in the current study, only three of the 13 individuals who were carrying *de novo* CNVs had an estimated IQ<70, with one further individual who had missing IQ data.

The identified CNVs affected a total of 262 genes. None of the genes spanned by the *de novo* CNVs have been highlighted by the only exome sequencing study of ADHD to date^3^ or implicated in ADHD based on a recent, large case-control GWAS^2^. However, several of the CNVs have been implicated by previously published ADHD case-control CNV association studies (which will include inherited and *de novo* CNVs); these include the robustly implicated 16p13.11 duplication and the 22q11.2 duplication^4–6^.

The overlap of *de novo* CNV loci in the regions 15q13.1-13.2, 16p13.11, 16p12.2 and 22q11.21 with loci previously implicated in ASD, DD/ID, Tourette disorder and schizophrenia is consistent with the notion that neuropsychiatric CNVs often have associations with a broad range of neurodevelopmental and psychiatric phenotypes^27^. Similar findings have emerged from the one sequencing study of ADHD, which highlighted strong overlap in the genes implicated in ADHD and ASD^3^, while GWAS demonstrate a moderate (r_g_=0.36) genetic correlation between ADHD and ASD^28^, and a previous CNV study suggested that the same biological pathways are impacted by CNVs in ADHD and ASD^29^. Our study adds to the growing body of literature supporting the relatively recent reconceptualization of ADHD as a neurodevelopmental disorder, as evidenced by its DSM-5 definition.

However while there is overlap in implicated CNV regions (see Table 2), the type of CNV is usually but not always the same. In particular, we observed a *de novo* 22q11 duplication and although such duplications have been observed to be associated with risk of other neurodevelopmental disorders, they are protective for schizophrenia^22^, while deletions at this locus (not observed here) are associated with schizophrenia as well as other neurodevelopmental impairments^21^.

Although ADHD is highly heritable, complex and polygenic, rare mutation discovery has been slower than for many other similarly heritable and polygenic disorders that include DD/ID, schizophrenia, ASD and Tourette disorder. For example, *de novo* CNV data are currently only available for around a total of 500 trios (Lionel and colleagues^16^ and the current study) and there have been no large, trio-based sequencing studies of ADHD. To identify associated variants, as well as damaging and likely causal rare mutations, will require very large sample sizes of ADHD that are not yet globally available.

In summary, we present findings which suggest that *de novo* CNVs likely contribute to ADHD risk and highlight the need for larger discovery studies for future biological insights.

## Data Availability

Genetic data are available via the PGC ADHD consortium.

https://www.med.unc.edu/pgc/results-and-downloads/adhd/?choice=Attention+Deficit+Hyperactivity+Disorder+%28ADHD%29

## Acknowledgements

The authors thank the families who participated in this project, the clinicians who supported it, the core lab staff in the MRC Centre for Neuropsychiatric Genetics and Genomics, as well as the National Centre for Mental Health (https://www.ncmh.info/). The work was supported by funding from the Wellcome Trust (Grants 079711 and 106047), Medical Research Council Centre (Grant No. MR/L010305/1), Health and Care Research Wales (grant number: 514032), Action Medical Research, and Baily Thomas.

## Disclosures

All of the authors report no conflicts of interest.

## Notes

### Competing Interest Statement

The authors have declared no competing interest.

